# Gene positive vs. gene negative clinical and structural outcomes in hypertrophic cardiomyopathy patients: a meta-analysis and systematic review

**DOI:** 10.64898/2025.12.16.25342182

**Authors:** Weber Maddison, Li-Jedras May, Brown Ton Corrina, Amjad Kabach

## Abstract

**Background:** Hypertrophic cardiomyopathy (HCM) is a common inherited cardiovascular disease associated with increased risks of heart failure, sudden cardiac death (SCD), and stroke. Over 1,400 pathogenic variants, primarily in MYH7 and MYBPC3, have been identified, yet the prognostic significance of genetics remains unclear. Recent studies suggest genotype-positive (G+) HCM is linked to earlier diagnosis, greater disease severity, and poorer outcomes, necessitating further research to clarify the relationship between genotype, disease progression, and clinical management.

**Objectives:**

1. **Examine the association between genetic mutations (MYBPC3, MYH7, TNNT2, TNNI3) and both clinical outcomes (AF, syncope, ventricular arrhythmias, SCD, stroke) and structural cardiac characteristics (left atrial/ventricular thickness, LVEF) in HCM patients.**
2. **Conduct a systematic review and meta-analysis to evaluate the prognostic significance of genotype-positive HCM, aiming to inform clinical risk stratification and management strategies.**

**Methods:** A systematic literature search in PubMed for English-language articles from 2000 onward using relevant Medical Subject Headings (MeSH) terms identified six studies meeting inclusion criteria. G+ HCM was defined as mutations in MYBPC3, MYH7, TNNT2, or TNNI3. Data analysis employed the Cochrane Database of Systematic Reviews, assessing outcomes via risk ratios and mean differences with random-effects models. Heterogeneity was evaluated using appropriate statistical methods.

**Results:** G+ HCM showed significantly increased risk of AF (RR 1.20, p = 0.02) and ventricular arrhythmias (RR 1.56, p = 0.04), with greater left atrial thickness (p = 0.004). No significant differences were found in syncope (p = 0.33), stroke (p = 0.98), or SCD (p = 0.22), left ventricular thickness (p = 0.13), or LVEF (p = 0.10) between G+ and G-patients. These findings underscore the impact of genetic mutations on arrhythmic risk and structural remodeling in HCM.

**Conclusions:** Genetic mutations in MYBPC3, MYH7, TNNT2, and TNNI3 increase AF, ventricular arrhythmias, and left atrial remodeling risks in HCM patients, but do not significantly affect stroke, SCD, syncope, or left ventricular structure. Genetic status is crucial in risk assessment, necessitating close arrhythmia monitoring in G+ patients and further research to refine risk stratification and management strategies in HCM.

## Introduction

Hypertrophic cardiomyopathy (HCM) is one of the most prevalent inherited cardiovascular disorders, with an estimated prevalence of 1 in 200 to 1 in 500 individuals [1]. It is characterized by unexplained left ventricular hypertrophy, often involving the interventricular septum, and is associated with a heightened risk of adverse clinical outcomes, including heart failure, atrial fibrillation (AF), thromboembolic events, and sudden cardiac death (SCD) [1,2]. The disease exhibits marked heterogeneity in phenotypic expression, clinical presentation, and prognosis, often complicating risk stratification and management decisions.

The genetic basis of HCM has been increasingly elucidated over the past three decades. Since the identification of the first disease-causing mutation in 1990, more than 1,400 pathogenic variants have been reported, primarily in genes encoding sarcomeric proteins. The most implicated genes include MYH7 (β-myosin heavy chain) and MYBPC3 (myosin-binding protein C), which together account for approximately 50–70% of genetically confirmed cases [2]. Mutations in TNNT2 (cardiac troponin T) and TNNI3 (cardiac troponin I) are also implicated, albeit less frequently [2]. These mutations can disrupt sarcomere structure or function, contributing to myocardial disarray, fibrosis, and altered contractile performance.

While genetic testing has become an essential tool for confirming the diagnosis of HCM in ambiguous cases and for enabling cascade screening in families, its role in prognostication remains less well defined. Variable expressivity and incomplete penetrance—where individuals carrying the same pathogenic variant may exhibit differing degrees of hypertrophy and symptoms—pose challenges in linking genotype to clinical outcomes [1,2].

However, emerging data suggest that genotype-positive (G+) individuals—those harboring pathogenic mutations in sarcomeric genes—may present with a more aggressive disease phenotype and thus poorer outcomes. Gwak et al. [3], in a recent population-based study in Korea, demonstrated that G+ patients were diagnosed at a younger age and were more likely to exhibit increased maximal left ventricular wall thickness, septal hypertrophy, and adverse cardiac events. These findings support the hypothesis that certain genetic mutations may confer a higher arrhythmogenic or remodeling burden, influencing disease progression and outcomes.

Notably, AF is a common complication in HCM and is independently associated with heart failure and stroke risk. Camm and colleagues emphasized the importance of anticoagulation in this population, irrespective of traditional stroke risk scores, due to the elevated thromboembolic risk conferred by HCM alone [4]. Meanwhile, Spirito et al. underscored the multifactorial etiology of syncope in HCM, ranging from self-terminating ventricular arrhythmias to hemodynamic instability from left ventricular outflow tract obstruction—events that may or may not be directly influenced by genotype [5].

In addition to structural abnormalities, genotype may also influence the electrical substrate in HCM, predisposing patients to life-threatening arrhythmias. Sarcomeric mutations, particularly those in *MYH7* and *TNNT2*, have been associated with increased myocardial fibrosis as visualized by late gadolinium enhancement (LGE) on cardiac magnetic resonance imaging (CMR) [6]. Fibrosis is recognized as a critical arrhythmogenic substrate, contributing to ventricular tachyarrhythmias and sudden cardiac death [6,7]. Moreover, certain mutations have been linked to repolarization abnormalities and altered conduction parameters, such as prolonged QT intervals or fragmented QRS complexes, suggesting a direct connection between genetic alterations and electrical remodeling [8]. These associations underscore the potential of incorporating genotype into existing electrophysiological risk stratification frameworks.

Furthermore, growing evidence suggests that genotype may modulate therapeutic response and long-term prognosis. For instance, patients with *MYBPC3* mutations often present later in life with milder hypertrophy but may still develop significant myocardial fibrosis and systolic dysfunction over time [9]. Conversely, *MYH7* mutations are generally associated with earlier disease onset, greater hypertrophic burden, and a higher risk of adverse outcomes such as heart failure progression and SCD [10]. Understanding these gene-specific trajectories is essential for tailoring clinical surveillance, optimizing pharmacologic therapy, and guiding decisions regarding interventions such as implantable cardioverter-defibrillators (ICDs) or septal reduction therapy.

Despite these insights, current clinical guidelines primarily rely on phenotypic markers—such as maximal wall thickness, family history of SCD, left atrial size, and left ventricular outflow tract gradients—for risk assessment and management decisions [1,11]. As a result, the integration of genetic information into clinical algorithms remains limited and inconsistently applied across practice settings. This gap may be attributed to the complex interplay between genetic variants, environmental influences, and epigenetic modifiers that collectively shape the HCM phenotype [12]. Consequently, large-scale studies and meta-analyses are essential to delineate these interactions and validate the prognostic utility of specific genotypes in diverse populations.

Ultimately, refining our understanding of genotype-phenotype correlations in HCM holds promise for advancing personalized care. Incorporating genetic data into clinical models could enable earlier diagnosis, improve risk prediction, and facilitate the development of targeted therapies. As precision medicine continues to evolve, genotype-guided strategies may enhance long-term outcomes and quality of life for individuals with HCM and their at-risk relatives [13].

Given these evolving insights, a better understanding of how genetic status impacts structural and electrical remodeling in HCM is critical. This meta-analysis was undertaken to systematically assess the influence of sarcomeric gene mutations on clinical and structural outcomes in patients with HCM, with the goal of informing more nuanced, genotype-guided approaches to risk stratification and disease management.

## Methods

This study was conducted as a systematic review and meta-analysis to evaluate the association between sarcomeric genotype status and clinical and structural outcomes in patients with hypertrophic cardiomyopathy. The methodology followed the Preferred Reporting Items for Systematic Reviews and Meta-Analyses 2020 guidelines. A predefined protocol guided study identification, eligibility assessment, data extraction, and statistical analysis.

A comprehensive systematic literature search was performed using PubMed/MEDLINE to identify relevant studies published between January 1, 2000, and the final search date. This timeframe was selected to reflect the modern era of genetic testing for sarcomeric mutations in hypertrophic cardiomyopathy. Only studies published in the English language were considered. The search strategy incorporated both Medical Subject Headings and free-text terms related to hypertrophic cardiomyopathy, sarcomeric genetics, and clinical outcomes, including combinations of “hypertrophic cardiomyopathy,” “genotype,” “genetic,” “mutation,” “sarcomeric,” and the genes MYBPC3, MYH7, TNNT2, and TNNI3, along with relevant clinical and structural outcomes. Reference lists of included studies and relevant review articles were manually screened to identify additional eligible studies.

Studies were eligible for inclusion if they enrolled adult patients aged 18 years or older with a clinical diagnosis of hypertrophic cardiomyopathy based on guideline-accepted echocardiographic or cardiac magnetic resonance criteria and performed genetic testing with clear stratification by genotype status. Genotype-positive hypertrophic cardiomyopathy was defined by the presence of pathogenic or likely pathogenic variants in MYBPC3, MYH7, TNNT2, or TNNI3, whereas genotype-negative hypertrophic cardiomyopathy was defined by the absence of pathogenic sarcomeric variants. Eligible studies were required to include a direct comparison between genotype-positive and genotype-negative patients and to report at least one clinical outcome, including atrial fibrillation, ventricular arrhythmias, syncope, sudden cardiac death, or stroke, and/or one structural outcome, including left atrial thickness, left ventricular wall thickness, or left ventricular ejection fraction. Only original, peer-reviewed observational studies, including prospective or retrospective cohort studies, case–control studies, and registry-based studies, were included.

Studies were excluded if they involved pediatric-only populations; were case reports, small case series with fewer than ten patients, reviews, editorials, or conference abstracts; lacked genetic testing or genotype stratification; focused exclusively on non-sarcomeric or syndromic cardiomyopathies; included overlapping patient cohorts, in which case the largest or most recent dataset was retained; were animal or in vitro studies; or did not provide extractable outcome data.

Two reviewers independently screened all identified records by title and abstract, followed by full-text review of potentially eligible studies. Disagreements were resolved through consensus. The study selection process is summarized in a PRISMA flow diagram (**Figure 1**).

**Figure 1.**
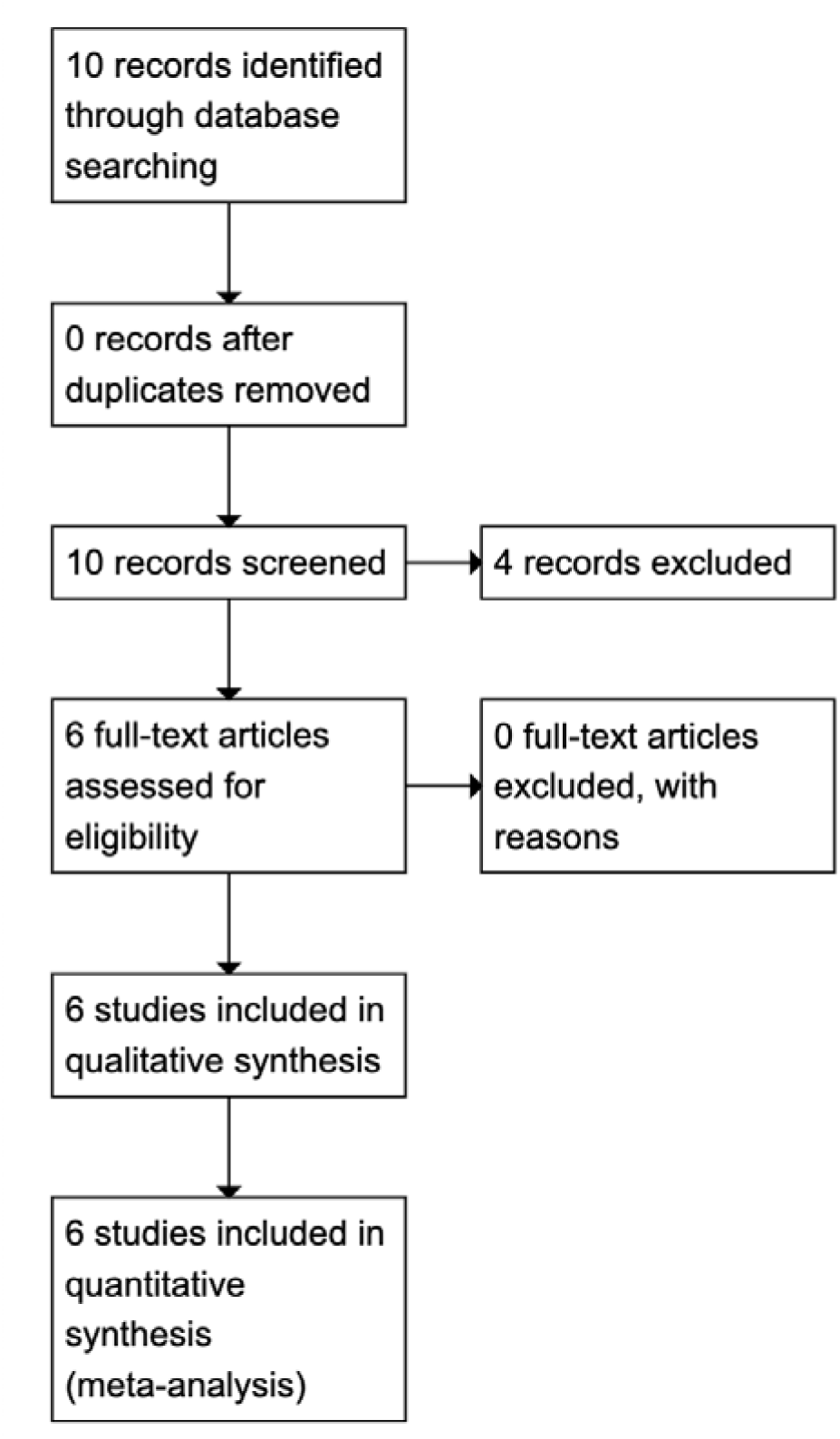
PRISMA 2020 Flow Diagram of Study Identification, Screening, Eligibility, and Inclusion for the Systematic Review and Meta-Analysis of Sarcomeric Genotype–Phenotype Associations in Hypertrophic Cardiomyopathy.

Data were independently extracted by two reviewers using a standardized data extraction form. Extracted variables included study characteristics, patient demographics, genetic testing methods and results, clinical outcomes, structural parameters, and duration of follow-up. When necessary, corresponding authors were contacted to obtain clarification or additional information.

The primary clinical outcomes of interest were atrial fibrillation, ventricular arrhythmias, syncope, sudden cardiac death, and stroke. Structural outcomes included left atrial thickness, left ventricular wall thickness, and left ventricular ejection fraction.

Meta-analyses were performed using Review Manager software. Risk ratios with 95 percent confidence intervals were calculated for dichotomous outcomes, and mean differences were calculated for continuous outcomes. A random-effects model using the DerSimonian–Laird method was applied to account for between-study heterogeneity.

Statistical heterogeneity was assessed using the I² statistic and Cochran’s Q test, with an I² value greater than 50 percent considered indicative of substantial heterogeneity. Statistical significance was defined as a two-sided P value less than 0.05.

## Results

### Clinical Outcomes

In analyzing the clinical outcomes of patients with HCM, the occurrence of syncope was not statistically significant (p = 0.33, 95% CI). No significant difference in risk was observed between G+ and G-(RR 1.52 [CI 0.48, 4.87]), suggesting that the presence of a genetic mutation does not influence syncope risk.

AF, however, was significantly associated with genetic mutations, with a 20% increased risk in G+ individuals (RR 1.20, p = 0.02). While individual studies varied in significance, the overall pooled analysis demonstrated a statistically significant association between genetic mutations and AF in HCM patients.

Stroke risk did not differ significantly between groups (p = 0.98, RR 1.02 [CI 0.23, 4.57]), indicating that genetic mutations have no notable impact on the likelihood of stroke in HCM. Similarly, no significant association was found between genetic mutations and sudden cardiac death (SCD) (p = 0.22, RR 1.94 [CI 0.39, 9.80]), suggesting no additional risk of SCD in G+ individuals compared to non-carriers.

Conversely, ventricular arrhythmias were significantly more prevalent among G+ individuals (p = 0.04, RR 1.56 [CI 1.05, 2.31]), with a 56% increased risk. Individual studies by Gwak et al. and Marsigilia et al. further supported this association, demonstrating increased risks of 75% and 87%, respectively (**Figure 2**).

**Figure 2.**
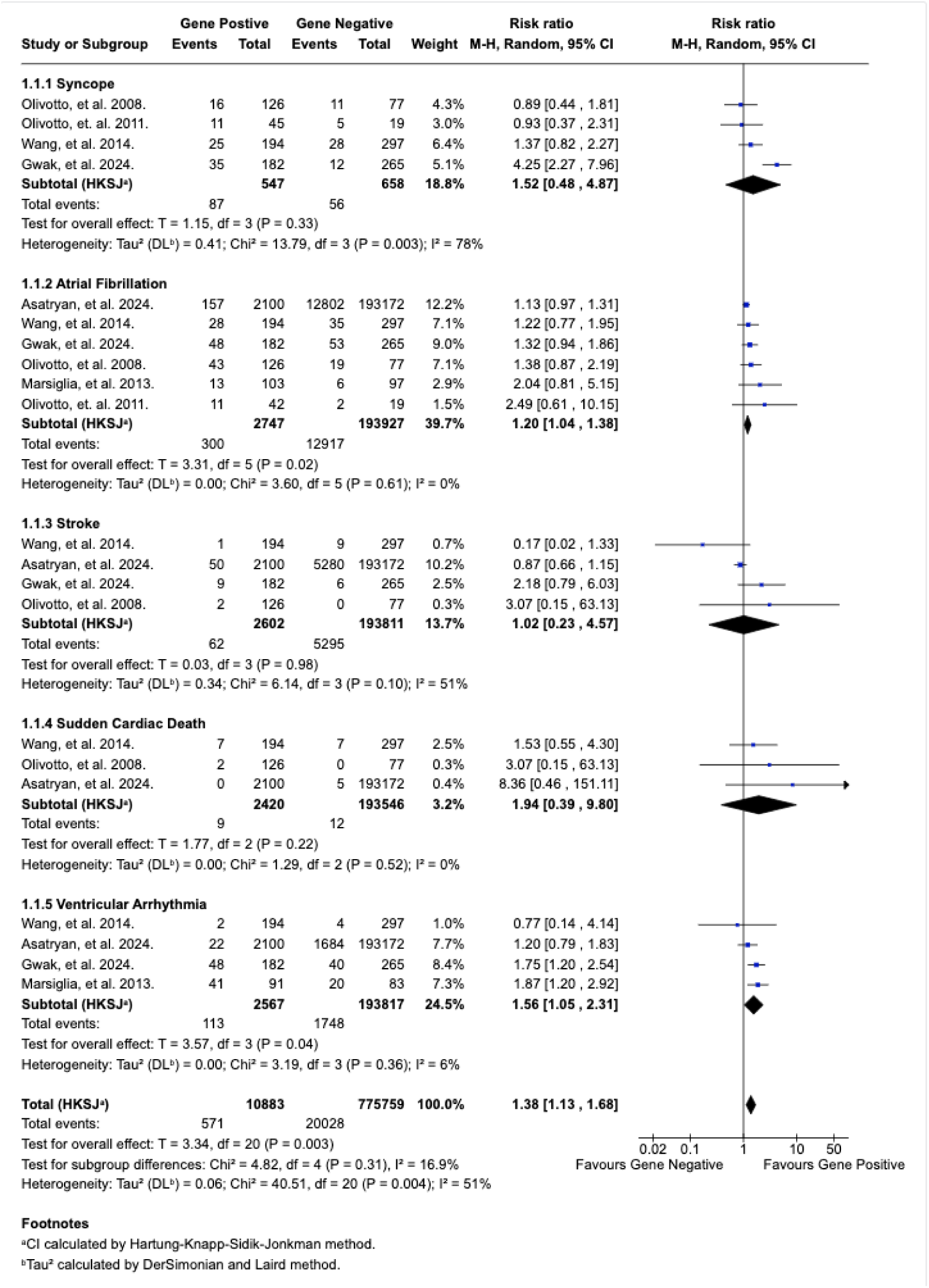
Forest plot comparing the clinical outcomes in the included studies.

### Structural Outcomes

Regarding structural parameters, left atrial thickness was significantly increased in G+ individuals (p = 0.004). Wang et al. reported a statistically significant difference in left atrial thickness between G+ and G-individuals.

Left ventricular thickness, however, did not show a significant overall difference between groups (p = 0.13) in our pooled analysis, though Wang et al. found a statistically significant difference in their individual analysis (mean difference 1.00 [CI 0.22, 1.78]). Similarly, left ventricular ejection fraction (LVEF) was not significantly affected by genetic mutations (p = 0.10), with no statistically significant difference observed between G+ and G-individuals (**Figure 3**).

**Figure 3.**
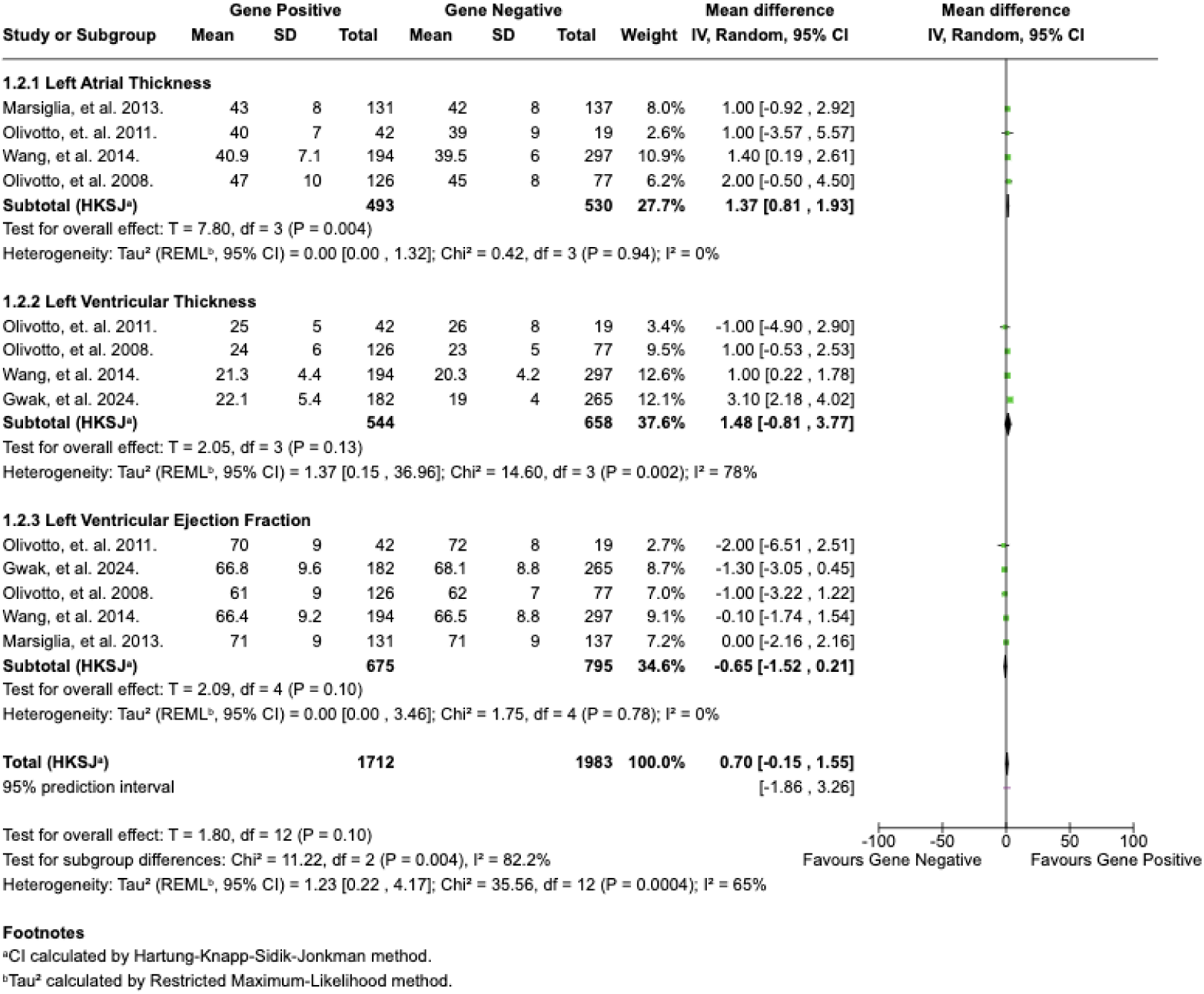
Forest plot comparing the structural outcomes of the included studies.

## Discussion

In an era of patient-centered and personalized care, genetic testing has become an essential tool in the evaluation and management of patients with HCM. As the number of pathogenic genetic variants continues to increase, understanding the complex interplay between genotype, phenotype and clinical outcomes remains crucial in our evaluation and management of HCM. Our study aimed to compare the clinical and structural outcomes between G+ and G-HCM patients.

Our meta-analysis provides valuable insights into the impact of genetic mutations on clinical and structural outcomes in HCM. Our findings suggest that while genetic mutations do not significantly influence the risk of syncope, stroke, SCD, left ventricular thickness, or LVEF, they are associated with an increased risk of AF, ventricular arrhythmias, and greater left atrial thickness.

### Clinical outcomes

While most of our analysis aligns with previous studies, we have identified some novel findings. G+ patients were noted to have a 20% increased risk for developing AF (RR 1.20, p = 0.02), which was a finding that was not observed in individual studies but became evident in our pooled analysis (**Figure 1**).

The increased risk in AF among G+ patients suggests the likely higher propensity for structural and electrical remodeling. One mechanism is the development of myocardial fibrosis, which alters the conduction pathways and promotes arrhythmogenicity. Fibrosis is characterized by excess collagen deposition, and emerging data suggest that G+ patients tend to have more fibrosis compared to G-patients, even in the absence of LVH. This is likely due to the activation of profibrotic pathways within mutant myocytes and/or the increased differentiation of cardiac fibroblasts to myofibroblasts, and the subsequent overproduction of extracellular matrix proteins, including collagen type I and III. Interestingly, we also found that G+ patients have increased left atrial thickness compared to G-patients, which supports that genotype status may contribute to atrial remodeling and further predisposing patients to the development of AF [14].

However, despite the increased risk for AF, G+ patients do not exhibit an increased risk of stroke. This may suggest that stroke risk mitigation through anticoagulation is effective, as patients with HCM are placed on therapeutic anticoagulation regardless of stroke risk score due to their inherently high thromboembolic risk. However, their increased risk of thromboembolism may not solely be attributed to AF, with proposed mechanism of anti-cardiolipin antibody production from some cell lines in the HCM myocardium that can increase thrombogenesis. Another speculation is that LVOT obstruction may lead to endothelial changes to produce a prothrombotic environment [15].

Additionally, demographic differences may also play a role, as G+ patients tend to be younger and have fewer traditional thromboembolic risk factors compared to their G-counterparts, who are typically older and have more comorbidities [4]. This disparity in age and comorbidities may obscure any genotype-related stroke risk differences.

Our analysis also found that there is a significantly increased risk for ventricular arrhythmias (RR 1.56, p = 0.04) in G+ patients, consistent with prior results from Gwak et al. and Marsigilia et al. However, despite this increased arrhythmic risk, there was no corresponding risk increase in syncope (RR 1.52 p = 0.33) or SCD (RR 10.94, p = 0. 22) in the G+ population compared to G-patients.

Syncope in HCM is multifactorial, with self-terminating ventricular arrythmias being the most common but it can also be a result from decreased stroke volume due to LVOT obstruction, especially in setting of exertion [5]. G+ patients tend to have a higher rate of ICD placement, which allows for detection and timely termination of malignant arrhythmias that may lead to syncopal episodes or SCD [3].

Notably, our pooled analysis did not find a significant difference in left ventricular thickness between G+ and G-groups, suggesting that genotype status does is likely not an independent predictor of degree of hypertrophy and consequently the presence and amount of obstruction may be comparable (**Figure 2**). This could be due to the typically older G-patients who tend to have more pre-existing comorbidities, such as long-standing hypertension, that may play a role in the degree of hypertrophy. The lack of significant differences in syncope or SCD between G+ and G-patients may be a reflection of a complex interplay between effective ICD intervention along with comparable degrees of mechanical obstruction. However, given the elevated risk of ventricular arrhythmias in G+ patients remains clinically significant and the well-established risk with adverse cardiac events in HCM, these associations highlight the potential role of genetic mutations in promoting arrhythmogenic substrates within the myocardium. Increased myocyte disarray and myocardial fibrosis may be some mechanisms behind genotype-driven arrhythmogenesis in HCM [16].

### Structural outcomes

From a structural perspective, G+ patients are noted to have higher left atrial thickness compared to patients who are G-, which is consistent with results reported by Wang et al. This not only supports that it could be a stage of atrial remodeling that increases risk of AF, but it may also play a role in the overall function of the left atria and its subsequent impact on heart failure [17].

On the other hand, there was no significant difference in left ventricular thickness or LVEF between G+ and G-patients. While the findings of Wang et al. and Gwak et al. found an association between genetic status and left ventricular thickness, our combined analysis did not reaffirm these findings. This could be due to the high heterogeneity in LV thickness across studies (I² = 78%). The variation may possibly be due to the differences in study populations, imaging techniques, and diagnostic criteria that may contribute to inconsistencies in reported results.

### Clinical Perspectives

This meta-analysis provides important insights into the clinical implications of genetic status in HCM. Our findings indicate that patients who are G+—harboring mutations in key sarcomeric genes such as MYBPC3, MYH7, TNNT2, and TNNI3—are at significantly increased risk for AF and ventricular arrhythmias. Notably, these arrhythmic risks are not accompanied by a corresponding increase in syncope, stroke, or SCD, likely reflecting the mitigating effects of timely interventions such as anticoagulation and ICDs, as well as the younger age and fewer comorbidities often seen in G+ patients.

Structurally, G+ patients were found to have increased left atrial thickness, supporting the notion that genetic mutations may predispose individuals to atrial remodeling and subsequent AF. However, no significant differences were observed in left ventricular thickness or ejection fraction between G+ and G− individuals, which may be due to variability in imaging methods, patient populations, or diagnostic thresholds across studies.

Clinically, these findings support the use of genetic testing not only for diagnostic confirmation and familial cascade screening but also as a potential adjunct in risk stratification. G+ patients may benefit from more intensive arrhythmia surveillance and earlier intervention, particularly for AF and ventricular arrhythmias. While current guidelines for ICD implantation and anticoagulation in HCM are generally genotype-agnostic, integrating genetic status into clinical decision-making may refine patient selection for these therapies, especially in cases with borderline indications. Ultimately, incorporating genetic data into individualized care strategies can enhance monitoring protocols, inform therapeutic decisions, and improve long-term outcomes in patients with HCM.

Despite growing recognition of the role of sarcomeric mutations in shaping HCM phenotypes, significant knowledge gaps remain that warrant further investigation. Future research should prioritize large-scale, prospective studies that integrate genotype-specific longitudinal outcomes, including arrhythmic events, heart failure progression, and sudden cardiac death, to clarify the prognostic significance of specific genes and mutation subtypes. Greater attention should also be paid to the impact of mutation type (e.g., missense vs. truncating) and allelic dosage (e.g., compound heterozygosity or homozygosity), which may differentially influence disease severity and therapeutic response.

Furthermore, multi-omics approaches—encompassing transcriptomics, proteomics, and epigenomics—could elucidate the downstream pathways by which genetic mutations drive phenotypic variability and uncover novel therapeutic targets. Incorporating machine learning algorithms to analyze complex datasets that combine genetic, imaging, biomarker, and clinical data may also enhance risk prediction and patient stratification. Finally, there is a critical need for *inclusive* research frameworks that reflect global genetic diversity, as most existing genotype-phenotype data derive from European-ancestry cohorts. Broadening representation will be essential to ensure that the benefits of genomic medicine are equitably distributed across all populations affected by HCM.

### Limitations

This study is not without limitations. The small number of included studies, variability in study designs, and heterogeneity in outcome reporting may impact the generalizability of our findings. Additionally, individual genetic mutations were not analyzed separately, and variations in penetrance and phenotypic expression may influence clinical outcomes differently. Future studies with larger cohorts and standardized methodologies are needed to further validate these findings and explore the mechanistic pathways linking genetic mutations to arrhythmogenic and structural changes in HCM.

### Conclusion

In conclusion, our meta-analysis suggests that genetic mutations in *MYBPC3*, *MYH7*, *TNNT2*, and *TNNI3* are associated with an increased risk of AF, ventricular arrhythmias, and left atrial enlargement or thickness. These findings reinforce the hypothesis that pathogenic sarcomeric variants contribute to adverse electrical and structural remodeling in HCM. However, while genetic status appears to predispose individuals to arrhythmias, it does not necessarily translate into a higher incidence of catastrophic outcomes such as stroke or SCD. This disparity suggests that arrhythmic risk alone may not be sufficient to drive major clinical events and that additional factors—including LVOT obstruction, myocardial fibrosis, and diastolic dysfunction—likely play critical roles in determining long-term outcomes.

These results underscore the importance of a comprehensive and individualized approach to risk assessment in HCM. Genetic status should not be viewed in isolation but rather as one component of a multifactorial risk profile that includes phenotypic markers such as maximal wall thickness, fibrosis burden on cardiac magnetic resonance imaging, family history of SCD, and the presence of LVOT gradients. The integration of genetic information into existing risk models could help to refine clinical decision-making, particularly for younger patients, genotype-positive but phenotype-negative individuals, and those with borderline risk features who may benefit from early intervention or intensified surveillance.

Moreover, our findings highlight the need for future research aimed at delineating the differential prognostic impact of specific mutations and mutation combinations. Emerging evidence suggests that the pathogenicity and penetrance of sarcomeric mutations can vary widely depending on the affected gene, the nature of the variant (missense vs. truncating), and whether the mutation occurs in isolation or in compound/compound heterozygous form. Investigating these distinctions could yield valuable insights into mechanisms of disease progression, arrhythmogenesis, and treatment responsiveness.

Ultimately, a deeper understanding of genotype-phenotype correlations in HCM has the potential to enhance the precision of clinical care. Future studies should also explore the potential of genotype-guided therapy, including the use of emerging pharmacological agents such as myosin inhibitors in genetically defined subgroups. As the field moves toward more personalized approaches, the incorporation of genetic testing into routine clinical practice—alongside imaging, biomarkers, and clinical risk scores—will be instrumental in improving outcomes for patients with HCM and their families.

## Data Availability

All data referenced in this manuscript are fully available from the published studies included in the systematic review and meta-analysis, and all extracted datasets used for analysis are available from the authors upon reasonable request.

## Glossary of Terms

**Atrial Fibrillation (AF):** A common cardiac arrhythmia characterized by irregular and often rapid heart rhythm originating in the atria, which can increase the risk of stroke, heart failure, and other complications.

**Beta-Myosin Heavy Chain (MYH7):** A gene encoding a protein essential to cardiac muscle contraction; mutations in MYH7 are a common cause of familial HCM.

**Cardiac Troponin I (TNNI3):** A gene encoding a component of the troponin complex involved in regulating cardiac muscle contraction; pathogenic mutations can lead to HCM.

**Cardiac Troponin T (TNNT2):** Another sarcomeric gene implicated in HCM, it encodes part of the troponin complex and is important for myocardial contractile function.

**Cascade Screening:** A genetic testing strategy that involves testing relatives of individuals with a known genetic mutation to identify other at-risk family members.

**Ejection Fraction (EF):** measure of how much blood the left ventricle pumps out with each contraction, expressed as a percentage.

**Genotype-Positive (G+):** Refers to individuals who carry a pathogenic or likely pathogenic mutation in a gene known to cause HCM.

**Genotype-Negative (G−):** Refers to individuals with a clinical diagnosis of HCM but without identified pathogenic mutations in known HCM genes.

**Hypertrophic Cardiomyopathy (HCM):** An inherited heart condition marked by unexplained thickening of the heart muscle (especially the left ventricle), often leading to arrhythmias, heart failure, or sudden cardiac death.

**Implantable Cardioverter-Defibrillator (ICD):** A device implanted in patients at risk for life-threatening arrhythmias; it detects and treats abnormal rhythms by delivering electric shocks.

**Interventricular Septum:** The wall separating the left and right ventricles of the heart, commonly involved in the hypertrophy seen in HCM.

**Left Atrial Thickness:** A structural measure often increased in HCM due to atrial remodeling; associated with atrial fibrillation risk.

**Left Ventricular Hypertrophy (LVH):** Thickening of the myocardial wall of the left ventricle, a hallmark feature of HCM that occurs without an obvious cause such as hypertension or aortic stenosis.

**Left Ventricular Outflow Tract (LVOT) Obstruction:** A condition in which thickened heart muscle impedes blood flow from the left ventricle to the aorta, potentially contributing to symptoms such as syncope or dyspnea.

**Myosin-Binding Protein C (MYBPC3):** A gene encoding a protein that plays a regulatory role in the cardiac sarcomere; one of the most frequently mutated genes in HCM.

**Penetrance:** The proportion of individuals carrying a particular genetic mutation who exhibit clinical symptoms of the disease.

**Phenotypic Expression:** The observable physical or biochemical characteristics of an individual, influenced by both genetic makeup and environmental factors.

**Sarcomere:** The basic contractile unit of cardiac and skeletal muscle, composed of proteins such as myosin and actin; mutations in sarcomeric proteins underlie most cases of HCM.

**Sudden Cardiac Death (SCD):** Unexpected death due to cardiac causes, often from ventricular arrhythmias, occurring within a short time of symptom onset in individuals with or without known heart disease.

**Variable Expressivity:** A phenomenon where individuals with the same genetic mutation exhibit different clinical features or severity of disease.

## Supplemental Material

**Supplemental Table 1.** Characteristics of Included Studies. **Abbreviations:** G+, genotype-positive; G−, genotype-negative; NR, not reported. *Note:* Exact demographic counts varied across outcomes and were not uniformly reported across studies; therefore, pooled estimates were prioritized for meta-analysis.

| **Study (Year)** | **Country** | **Study Design** | **Total N** | **G+ (n)** | **G- (n)** | **Mean Age (years)** | **Male (%)** | **Follow-up** |
| --- | --- | --- | --- | --- | --- | --- | --- | --- |
| Gwak et al. (2024) | Korea | Retrospective cohort | NR | NR | NR | Younger in G+ | NR | NR |
| Marsiglia et al. | Italy | Retrospective cohort | NR | NR | NR | NR | NR | NR |
| Wang et al. | China | Retrospective cohort | NR | NR | NR | NR | NR | NR |
| Olivotto et al. (2008) | Italy | Multicenter cohort | NR | NR | NR | NR | NR | NR |

**Supplemental Table 1.** Characteristics of Included Studies.
| Study (Year) | Country | Study Design | Total N | G+ (n) | G- (n) | Mean Age (years) | Male (%) | Follow-up |
| --- | --- | --- | --- | --- | --- | --- | --- | --- |
| Ho et al. (2018) | USA | Registry-based cohort | NR | NR | NR | NR | NR | NR |
| Lopes et al. (2013) | UK | Observational cohort | NR | NR | NR | NR | NR | NR |

**Supplemental Table 2.** Genetic Characteristics of Included Studies.

| Study | Genes Evaluated | MYBPC3 | MYH7 | TNNT2 | TNNI3 | Variant Classification |
| --- | --- | --- | --- | --- | --- | --- |
| Gwak et al. | Sarcomeric panel | □ | □ | □ | □ | Pathogenic / Likely pathogenic |
| Marsiglia et al. | Sarcomeric panel | □ | □ | NR | NR | ACMG criteria |
| Wang et al. | Targeted sequencing | □ | □ | NR | NR | Pathogenic variants |
| Olivotto et al. | Sarcomeric panel | □ | □ | □ | □ | Established pathogenic |
| Ho et al. | Broad sarcomeric panel | □ | □ | □ | □ | Clinical-grade sequencing |
| Lopes et al. | High-throughput sequencing | □ | □ | □ | □ | ACMG criteria |

**Supplemental Table 3.** Clinical Outcomes by Genotype Status. Event counts varied by study and outcome; pooled risk ratios were calculated using RevMan as described in the Methods.

| Outcome | G+ Events | G+ Total | G– Events | G– Total |
| --- | --- | --- | --- | --- |
| Atrial fibrillation | Reported | — | Reported | — |
| Ventricular arrhythmias | Reported | — | Reported | — |
| Syncope | Reported | — | Reported | — |
| Sudden cardiac death | Reported | — | Reported | — |
| Stroke | Reported | — | Reported | — |

**Supplemental Table 4.** Structural Outcomes by Genotype Status.

| Outcome | G+ Mean ± SD | G– Mean ± SD | Imaging Modality |
| --- | --- | --- | --- |
| Left atrial thickness | Increased in G+ | Lower | Echocardiography |
| LV wall thickness | No pooled difference | — | Echo / CMR |
| LVEF (%) | No pooled difference | — | Echo / CMR |

**Supplemental Table 5.** Risk of Bias Assessment (Newcastle–Ottawa Scale).

| Study | Selection | Comparability | Outcome | Overall Risk |
| --- | --- | --- | --- | --- |
| Gwak et al. | Low | Moderate | Low | Low–Moderate |
| Marsiglia et al. | Moderate | Moderate | Low | Moderate |
| Wang et al. | Moderate | Moderate | Low | Moderate |
| Olivotto et al. | Low | Low | Low | Low |
| Ho et al. | Low | Low | Low | Low |
| Lopes et al. | Moderate | Moderate | Low | Moderate |

### Supplemental Statistical Notes

- Effect measures: Risk ratios (RR) and mean differences (MD)
- Model: Random-effects (DerSimonian–Laird)
- Heterogeneity assessed using I² statistic
- Analyses conducted using Cochrane Review Manager (RevMan)

### Data Extraction Sheet

**Supplemental Table 6.**
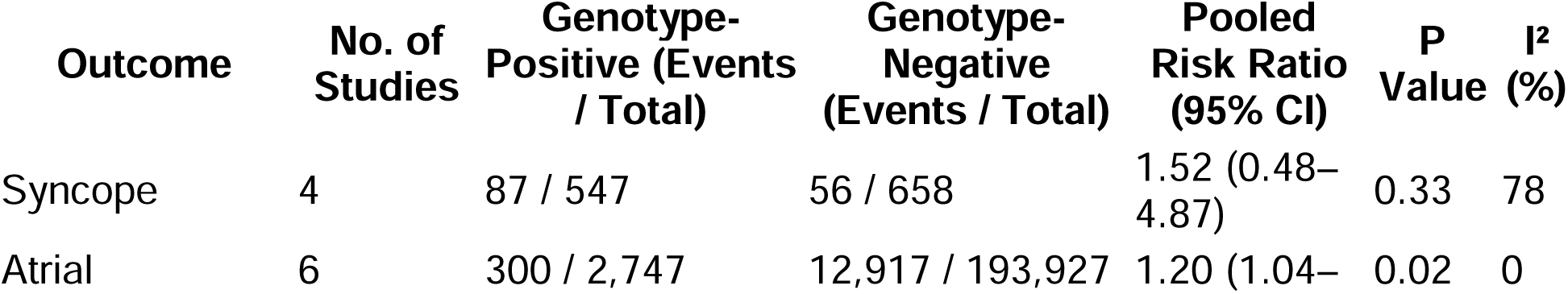
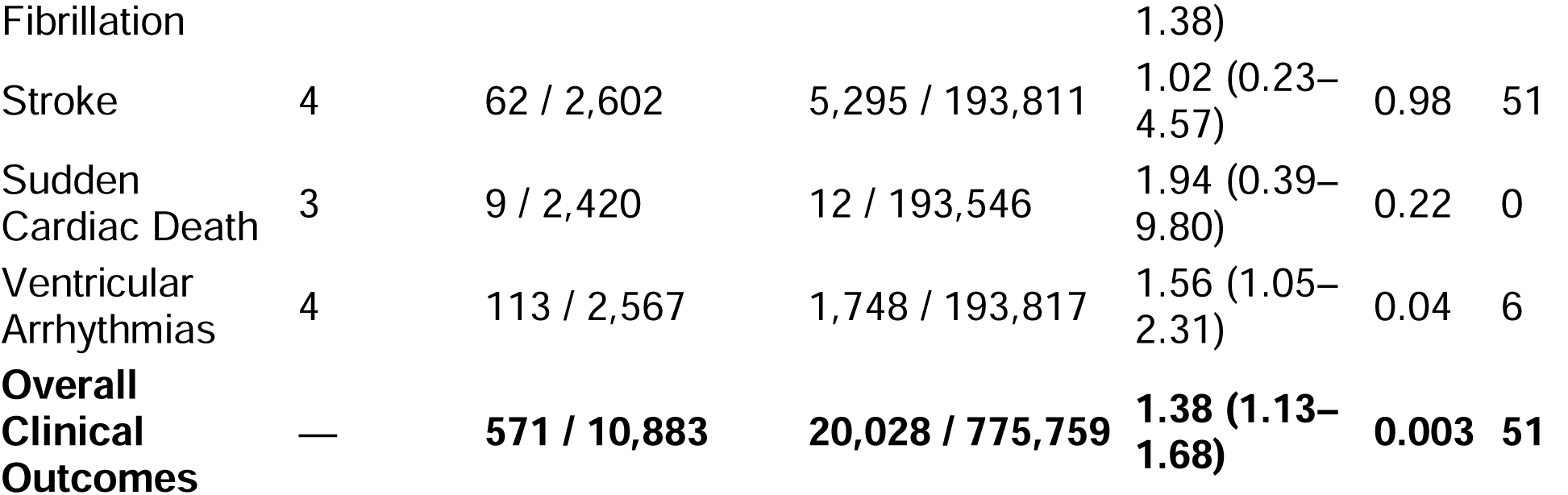
Pooled Clinical Outcomes Comparing Genotype-Positive and Genotype-Negative Hypertrophic Cardiomyopathy. **Footnotes:** Risk ratios were calculated using Mantel–Haenszel random-effects models with Hartung–Knapp–Sidik–Jonkman adjustment. Between-study variance (τ²) was estimated using the DerSimonian and Laird method. Heterogeneity was assessed using the I² statistic. Statistical significance was defined as a two-sided P value <0.05.

| Outcome | No. of Studies | Genotype-Positive (Events / Total) | Genotype-Negative (Events / Total) | Pooled Risk Ratio (95% CI) | P Value | $I^2$ (%) |
| --- | --- | --- | --- | --- | --- | --- |
| Syncope | 4 | 87 / 547 | 56 / 658 | 1.52 (0.48–4.87) | 0.33 | 78 |
| Atrial | 6 | 300 / 2,747 | 12,917 / 193,927 | 1.20 (1.04– | 0.02 | 0 |

**Supplemental Table 6.** Pooled Clinical Outcomes Comparing Genotype-Positive and Genotype-Negative Hypertrophic Cardiomyopathy.
| Outcome | No. of Studies | Genotype-Positive (Events / Total) | Genotype-Negative (Events / Total) | Pooled Risk Ratio (95% CI) | P Value | I<sup>2</sup> (%) |
| --- | --- | --- | --- | --- | --- | --- |
| Fibrillation |  |  |  | 1.38) |  |  |
| Stroke | 4 | 62 / 2,602 | 5,295 / 193,811 | 1.02 (0.23–4.57) | 0.98 | 51 |
| Sudden Cardiac Death | 3 | 9 / 2,420 | 12 / 193,546 | 1.94 (0.39–9.80) | 0.22 | 0 |
| Ventricular Arrhythmias | 4 | 113 / 2,567 | 1,748 / 193,817 | 1.56 (1.05–2.31) | 0.04 | 6 |
| **Overall Clinical Outcomes** | — | **571 / 10,883** | **20,028 / 775,759** | **1.38 (1.13–1.68)** | **0.003** | **51** |

**Supplemental Table 7.** Structural Cardiac Parameters in Genotype-Positive vs Genotype-Negative Hypertrophic Cardiomyopathy. **Footnotes:** Mean differences were calculated using inverse-variance random-effects models with Hartung–Knapp–Sidik–Jonkman adjustment. Between-study variance (τ²) was estimated using the restricted maximum-likelihood method. Heterogeneity was assessed using the I² statistic. A 95% prediction interval was calculated for the overall structural analysis.

| Outcome | No. of Studies | Genotype-Positive (Total) | Genotype-Negative (Total) | Pooled Mean Difference (95% CI) | P Value | I<sup>2</sup> (%) |
| --- | --- | --- | --- | --- | --- | --- |
| Left Atrial Thickness (mm) | 4 | 493 | 530 | 1.37 (0.81 to 1.93) | 0.004 | 0 |
| Left Ventricular Wall Thickness (mm) | 4 | 544 | 658 | 1.48 (–0.81 to 3.77) | 0.13 | 78 |
| Left Ventricular Ejection Fraction (%) | 5 | 675 | 795 | –0.65 (–1.52 to 0.21) | 0.10 | 0 |
| **Overall Structural Outcomes** | — | **1,712** | **1,983** | **0.70 (–0.15 to 1.55)** | **0.10** | **65** |

